# Bi-allelic variants in *CHKA* cause a neurodevelopmental disorder with epilepsy and microcephaly

**DOI:** 10.1101/2021.10.21.21265050

**Authors:** Chiara Klöckner, J. Pedro Fernandez Murray, Mahtab Tavasoli, Heinrich Sticht, Gisela Stoltenburg-Didinger, Leila Motlagh Scholle, Somayeh Bakhtiari, Michael C. Kruer, Hossein Darvish, Saghar Ghasemi Firouzabadi, Alex Pagnozzi, Anju Shukla, Katta Mohan Girisha, Dhanya Lakshmi Narayanan, Parneet Kaur, Reza Maroofian, Maha S. Zaki, Mahmoud M. Noureldeen, Andreas Merkenschlager, Janina Gburek-Augustat, Elisa Cali, Selina Banu, Kamrun Nahar, Stephanie Efthymiou, Henry Houlden, Rami Abou Jamra, Jason Williams, Christopher R. McMaster, Konrad Platzer

**Author notes:** Correspondence to: Konrad Platzer, Philipp-Rosenthal-Str. 55, 04103 Leipzig, Germany. Correspondence may also be addressed to Christopher R. McMaster, Department of Pharmacology, Dalhousie University, Halifax, Nova Scotia B3N 0A1, Canada.

## Abstract

The Kennedy pathways catalyze the *de novo* synthesis of phosphatidylcholine and phosphatidylethanolamine, the most abundant components of eukaryotic cell membranes. In recent years, these pathways have moved into clinical focus since four out of ten genes involved have been associated with a range of autosomal recessive rare diseases such as a neurodevelopmental disorder with muscular dystrophy (*CHKB*), bone abnormalities and cone-rod dystrophy (*PCYT1A*), and spastic paraplegia (*PCYT2, SELENOI*).

We identified six individuals from five families with bi-allelic variants in *CHKA* presenting with severe global developmental delay, epilepsy, movement disorders, and microcephaly. Using structural molecular modeling and functional testing of the variants in a in a cell-based *S. cerevisiae* model, we determined that these variants reduce the enzymatic activity of *CHKA* and confer a significant impairment of the first enzymatic step of the Kennedy pathway.

In summary, we present *CHKA* as a novel autosomal recessive gene for a neurodevelopmental disorder with epilepsy and microcephaly.

## Introduction

Eukaryotic membranes are dependent on the precise compositions of glycerophospholipids, the most abundant being PC and PE. PC and PE account for more than half of the phospholipid species in eukaryotic membranes and are synthesized *de novo* by the Kennedy pathways.^1^

*CHKA* encodes for choline kinase alpha, an enzyme that catalyzes the first step of phospholipid synthesis in the Kennedy pathway. Together with its paralog *CHKB*, it phosphorylates either choline or ethanolamine using ATP resulting in phosphocholine or phosphoethanolamine and ADP.^2,3^ Bi-allelic variants in *CHKB* are associated with a neurodevelopmental disorder with muscular dystrophy characterized by intellectual disability, microcephaly, hypotonia and structural mitochondrial abnormalities (MIM 602541).^4–6^ In recent years, variants in further genes involved in the Kennedy pathway have been described to cause recessive hereditary disorders, ranging from bone abnormalities with cone rod dystrophy (*PCYT1A*, MIM 608940)^7^ to neurodevelopmental disorders such as complex spastic paraplegia (*PCYT2*, MIM 618770; *SELENOI*, MIM 618768).^8,9^ Similar lipid metabolic pathways have also been associated with hereditary motor neuron degenerative diseases.^10^

In this study, we describe six individuals from five families with homozygous and compound-heterozygous pathogenic variants in *CHKA*. They present with a severe neurodevelopmental disorder characterized by DD/ID, epilepsy, and microcephaly. We also verified altered protein function using structural *in silico* modeling and functional testing of variants in a cell-based model.

## Materials and methods

### Standard protocol approvals

The study was approved by the ethics committee of the University of Leipzig, Germany (402/16-ek). All families provided informed consent for clinical testing and publication.

### Research cohort and identification of variants

By using matchmaking platforms and international collaborations, six individuals from five families harboring homozygous and compound-heterozygous variants in *CHKA* were identified.^11^ Phenotypic and genotypic information were obtained from the referring collaborators using a standardized questionnaire. Causality of both truncating and missense variants were assessed according to the guidelines of the ACMG (Table S1).^12^

For individuals 1.1 and 1.2, quattro ES for the parents and the two affected children were performed. Variants identified by ES were validated using Sanger sequencing. For individuals 2, 3 and 5 Singleton ES was performed. Validation of all variants identified by ES and bi-allelic segregation analysis were done by Sanger sequencing. For individual 4, trio ES was performed, and Sanger sequencing validated the identified variants and segregation analysis (for further details see Supplemental Methods 1).

### Subcloning of human *CHKA* allelic variants

The DNA for a wild type ORF of human *CHKA* C-terminally tagged with Myc and FLAG epitopes was amplified by PCR from Origene plasmid RC219747 using HiFi Platinum *Taq* polymerase and 5’-*Xba*I-*CHKA* and 3’-*Sal*I-*CHKA* primers. These primers introduce the mentioned restriction sites flanking a *CHKA*-Myc-FLAG ORF. A 1.4 kilobase DNA product was gel purified and subcloned into a pCR1.1TOPO vector rendering plasmids carrying the amplicon. Sequencing and restriction analyses revealed all plasmids tested carried a similar gap spanning around 200 bp from ∼ nt 100 to ∼ nt 320 of the *CHKA* ORF. Further attempts for PCR amplification under processing conditions over GC rich stretches using Phusion and Q4 DNA polymerases also failed to render a wild type *CHKA* ORF.

To rectify this, a sequenced gapped clone of *CHKA* was released from the pCR1.1TOPO-*CHKA* backbone by *Xba*I/*Sal*I digestion and subcloned into the yeast expression vector p416-GPD. To restore an intact wild type *CHKA* ORF into the yeast expression vector, a 423 bp *Blp*I/*BamH*I fragment that contains the unamplifiable *CHKA* GC rich region was isolated directly from the Origene RC219747 plasmid and subcloned into the p416-GPD vector containing the *CHKA* ORF missing the GC rich region. DNA sequencing confirmed that the *CHKA* ORF was now intact, full-length, and fused in frame to C-terminal myc and FLAG tags. The p.(Arg141Trp), p.(Pro194Ser), and p.(Phe341Leu) variants were generated by site directed mutagenesis on the p416-GPD-CHKA (missing the GC rich region) using the QuikChange mutagenesis kit (Agilent) following manufacturer’s instructions. Upon confirmation of introduction of the point mutations by DNA sequencing, the GC rich region was subcloned from a *Blp*I/*BamH*I fragment directly from the Origene RC219747 plasmid. DNA sequencing was used to confirm the ORF for each of these plasmids.

### Yeast transformation and cell culture

Wild type BY4742 and otherwise isogenic *cki1*Δ::KanMX6 strains were transformed with plasmid DNA following standard yeast protocols and selected on media for plasmid maintenance. Transformed cells were grown to logarithmic phase at 30°C in liquid medium enabling plasmid selection and retention.

### Protein extraction and western blot analysis

Logarithmic grown yeast cells were harvested, washed, and taken up in lysis buffer (50 mM Tris-HCl, 0.3 M sucrose, 1 X Complete protease inhibitor cocktail (Roche), 2 mg/ml pepstatin A, 1 mM PMSF) at 30 OD units/ml. Cells were broken by glass bead beating and supernatants of a 500 *g* x 5 min centrifugation were collected. Protein amount was determined by the Bradford method and equal amounts of protein were subjected to SDS-PAGE analysis followed by western blotting. Monoclonal antibodies against Myc were used to determine CHKA expression with yeast Pgk1 used as a loading control.

### Choline kinase activity

Choline kinase activity was estimated by the synthesis of phosphocholine from radiolabeled choline using yeast cytosolic fractions as sources of enzyme, followed by TLC to separate substrate from product. For cytosolic fraction preparation, logarithmic growing yeast cultures were chilled out in an ice-water bath, harvested, washed, and taken up in ice-cold lysis buffer (50 mM Tris-HCl, 0.3 M sucrose, 1 mM EDTA, 3 mM DTT, 2ug/ml pepstatin A) at 60 OD units/ml. Cells were broken by glass bead beating and the whole cell extract was subjected to 400,000 *g* centrifugation for 15 min at 4°C.

Choline kinase activity was measured using 2 mM [^14^C]choline (4,000 dpm/nmol), 10 mM ATP, 10 mM MgSO_4_ in 100 mM Glycine-NaOH pH 9.6 buffer containing 1.5 mM DTT. Enzyme reactions proceeded for 1 h at 37°C, stopped by heating at 96° C for 5 min, and centrifuged at 21,000 *g* for 10 min. Aliquots of the supernatants were loaded on silica gel TLC plates and chromatographed using a CH_3_OH:0.6 % NaCl:NH_4_OH (50:50:5) solvent system. TLC plates were radiometrically scanned and the radiolabelled band corresponding to phosphocholine was scraped into vials for liquid scintillation counting. Chromatographic positions of choline and phosphocholine were assigned based on the mobility of radiolabelled standards. Choline kinase activity was assayed under conditions of linear increase of product formation depending on reaction time and enzyme amount. Phosphocholine formation never exceeded 5 % of total substrate. The relative abundance of the different allelic variants of *CHKA* present in the high-speed supernatants were estimated by western blotting and used for normalizing choline kinase activity comparison across the variants. Data represent the mean +/-standard error of four determinations performed in triplicate.

### Skeletal muscle biopsy

Details on muscle biopsy and staining are available in the Supplemental Methods 2.

### Structural modeling

The structural effect of the variants was investigated based on the crystal structures of CHKA in complex with ADP (PDB:3G15^13^) or phosphocholine (PDB: 2CKQ^14^). Variants were modelled with SwissModel^15^ and RasMol^16^ was used for structure analysis and visualization.

### Data availability

The authors confirm that the data supporting the findings of this study are available within the article and its supplementary material.

## Results

### Clinical description

All five individuals aged between 2 and 11 years were affected by a neurodevelopmental disorder. First symptoms were noted in the first year of life and comprise severe DD/ID, seizures, and microcephaly. Further symptoms include movement disorders, and abnormal muscle tone. An overview of the clinical symptoms is presented in Table 1. Further clinical data and MRI images are presented in Supplemental case reports.

**Table 1.** Clinical information on individuals with bi-allelic variants in *CHKA*.

| Individual ID | 1.1 | 1.2 | 2 | 3 | 4 | 5 |
| --- | --- | --- | --- | --- | --- | --- |
| **Genomic position (NC_000011.9)** | g.67864527G>A | g.67864527G>A | g.67864527G>A | g.67842234G>A | g.67888631dup, g.67833357A>G | g.67888643A>G |
| **cDNA (NM_001277.2)** | c.421C>T | c.421C>T | c.421C>T | c.580C>T | c.14dup, c.1021T>C | c.2T>C |
| **Protein alteration (NP_001268.2)** | p.(Arg141Trp) | p.(Arg141Trp) | p.(Arg141Trp) | p.(Pro194Ser) | p.(Cys6Leufs*19), p.(Phe341Leu) | p.(Met1?) |
| **Zygosity** | Homozygous | Homozygous | Homozygous | Homozygous | Compound-heterozygous | Homozygous |
| **Consanguinity** | Yes | Yes | Yes | Yes | No | Yes |
| **Age at last assessment** | removed (medRxiv policy) | removed (medRxiv policy) | removed (medRxiv policy) | removed (medRxiv policy) | removed (medRxiv policy) | removed (medRxiv policy) |
| **Sex** | Male | Female | Male | Male | Male | Female |
| **Microcephaly** | Yes (42 cm, -7 SD) | Yes (45 cm, -4 SD) | Yes (44 cm, -6 SD) | Yes (41,8 cm, -5 SD) | Yes (49,6 cm, -3.3 SD) | Yes (44,5 cm, -6 SD) |
| **Short stature** | Yes | Yes | Yes (82 cm, - 4 SD) | No | No | Yes (106 cm, - 2.3 SD) |
| **Global developmental delay / Intellectual Disability** | Severe | Severe | Severe | Severe/profound | Severe | Severe/profound |
| **Gross motor delay** | No walking achieved | Assisted walking since age 3 years | No walking achieved | No walking achieved | No walking achieved | No walking achieved |
| **Speech and language** | No speech | No speech | No speech | No speech | No speech | No speech |
| **Seizures** |  |  |  |  |  |  |
| **Epileptic encephalopathy** | Yes | Yes | Yes | Yes | Yes | Yes |
| **Age at seizure onset** | Infancy | Infancy | < 5 years (medRxiv policy) | < 5 years (medRxiv policy) | < 5 years (medRxiv policy) | < 5 years (medRxiv policy) |
| **Seizure type at onset** | Epileptic spasms | Epileptic spasms | Epileptic spasms | Generalized seizures | Epileptic spasms | Generalized seizures |
| **Further seizure type** | Focal and generalized seizures | Focal and generalized seizures |  | Myoclonic seizures | Tonic-clonic seizures | Tonic seizures |
| **Brain MRI** | Normal | Normal | Normal | Deep white matter hypomyelination, thin corpus callosum, faint increased signal intensity in lentiform nucleus at T-FLAIR | Hypomyelination of occipital white matter, initially pronounced (age 15 months), normal MRI at follow-up (age 6 years) | Not done |
| **Movement Disorder** | None | None | None | Dyskinesia, rigidity | Dyskinesia | choreoathetotic movements |
| **Hyperreflexia** | Yes | Yes | Yes | Yes | Yes | Unknown |
| **Muscle tone** | Hypertonia | Hypertonia | Hypotonia | Hypotonia | Hypertonia | Hypotonia |
| **Additional Symptoms** | Scoliosis, nystagmus, aggressive behavior | Scoliosis, nystagmus, aggressive behavior | Hyperactivity, self injurious behaviour | Autistic behavior, nystagmus, high arched palate, poor visual acuity, cortical visual loss, diffuse RPE, moderate retinal dysfunction, dysmorphic facial features | Scoliosis, myopia, recurrent kidney stones | Aggressive behavior, poor sleep, feeding problems |

REP = retinal pigment epithelium

#### Individuals 1.1 and 1.2

This case report has been removed from the preprint version to comply with medrxiv policy. Please see the published version or contact the authors if you are interested in this information.

#### Individual 2

This case report has been removed from the preprint version to comply with medrxiv policy. Please see the published version or contact the authors if you are interested in this information.

#### Individual 3

This case report has been removed from the preprint version to comply with medrxiv policy. Please see the published version or contact the authors if you are interested in this information.

#### Individual 4

This case report has been removed from the preprint version to comply with medrxiv policy. Please see the published version or contact the authors if you are interested in this information.

#### Individual 5

This case report has been removed from the preprint version to comply with medrxiv policy. Please see the published version or contact the authors if you are interested in this information.

### Genotypic spectrum and structural modeling

Three different missense variants, one start-loss variant and one truncating variant have been observed in this cohort.

Individuals 1.1, 1.2 and 2 carry the homozygous variant p.(Arg141Trp). Individuals 1.1 and 1.2 are siblings. The CHKA structure indicates that Arg141 is in the vicinity of the ADP binding site and stabilizes the structure by forming hydrogen bonds to Pro130 and Thr133 (Fig. 1B_1). These interactions cannot be formed by the uncharged aromatic Trp141 sidechain in the variant (Fig. 1B_2) thereby causing destabilization close to the ADP binding site.

**Figure 1.**
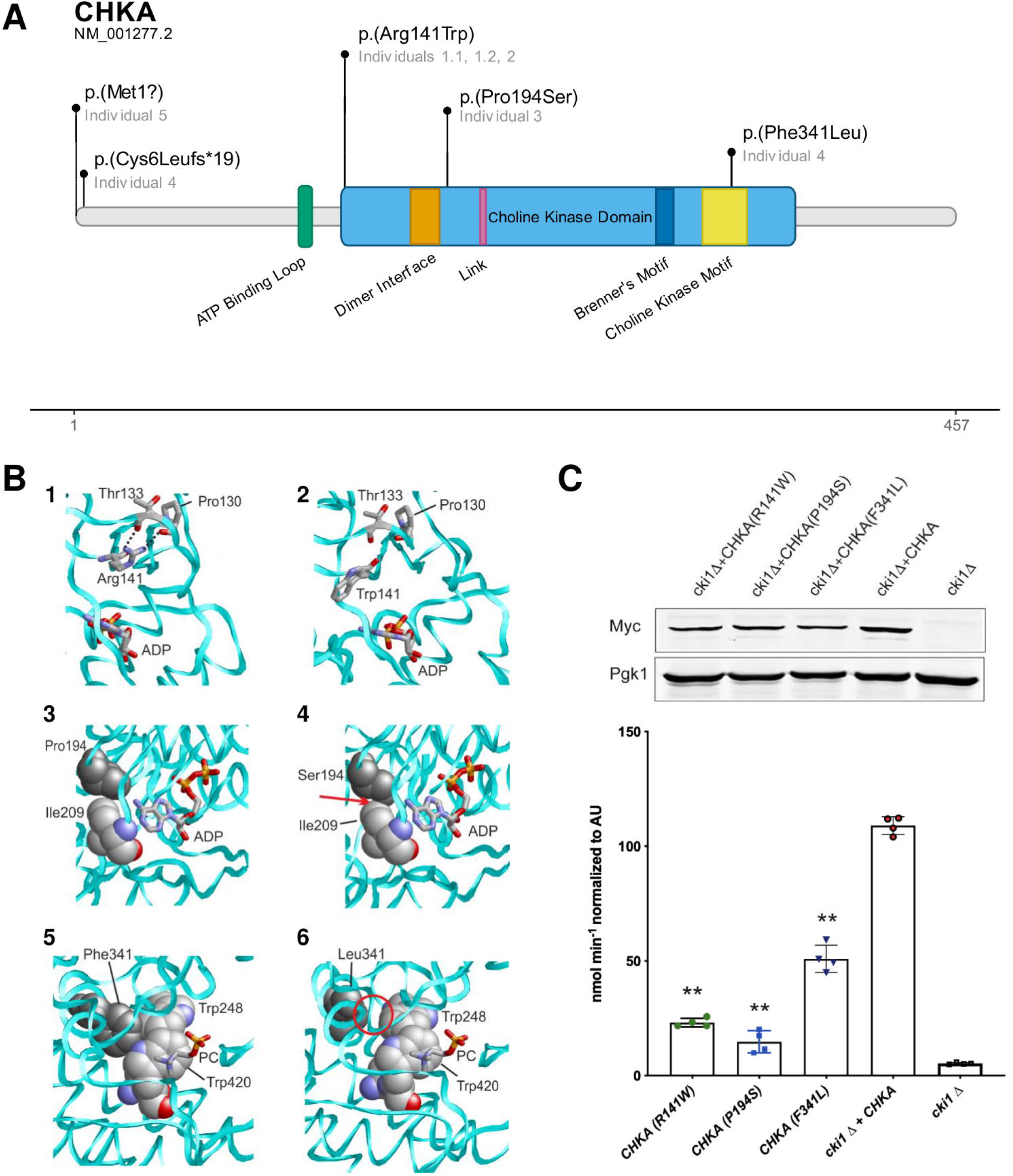
Overview on location of variants, structural modeling, and functional testing in an *S. cerevisiae* model. **(A)** Location of variants in *CHKA* with respect to reported domain structure.^14^ **(B)** Structural effect of *CHKA* sequence variants. (1) Arg141 forms stabilizing hydrogen bonds (black dotted lines) with Pro130 and Thr133 in the vicinity of the ADP binding site. Interacting residues and ADP are shown in stick presentation (atom-type coloring) and are labelled. The CSKH protein backbone is depicted as cyan ribbon. (2) Trp141 cannot form the stabilizing hydrogen bonds to Pro130/Thr133 and adopts a different sidechain orientation. Color coding as in (1). (3) Pro194 (grey) is located in a turn close to the ADP binding site. (4) The Ser194 sidechain causes steric problems (indicated as red arrow) with the adjacent Ile209 thereby destabilizing the structure. (5) Phe341 (grey) is part of a hydrophobic cluster close to the binding site of phosphocholine (PC; shown as sticks). Residues of the hydrophobic cluster are shown in space-filled presentation. (6) The presence of the nonaromatic Leu341 results in a loss of hydrophobic interactions (denoted as red circle). **(C)** Western blot of human CHKA expressed in yeast demonstrates that each allele was expressed at a similar level. Pgk1 is the loading control. Choline kinase activity of each allele, normalized to the level of CHKA expressed, was determined. It was determined that each patient-derived allele possessed reduced choline kinase activity.

**Figure 2.**
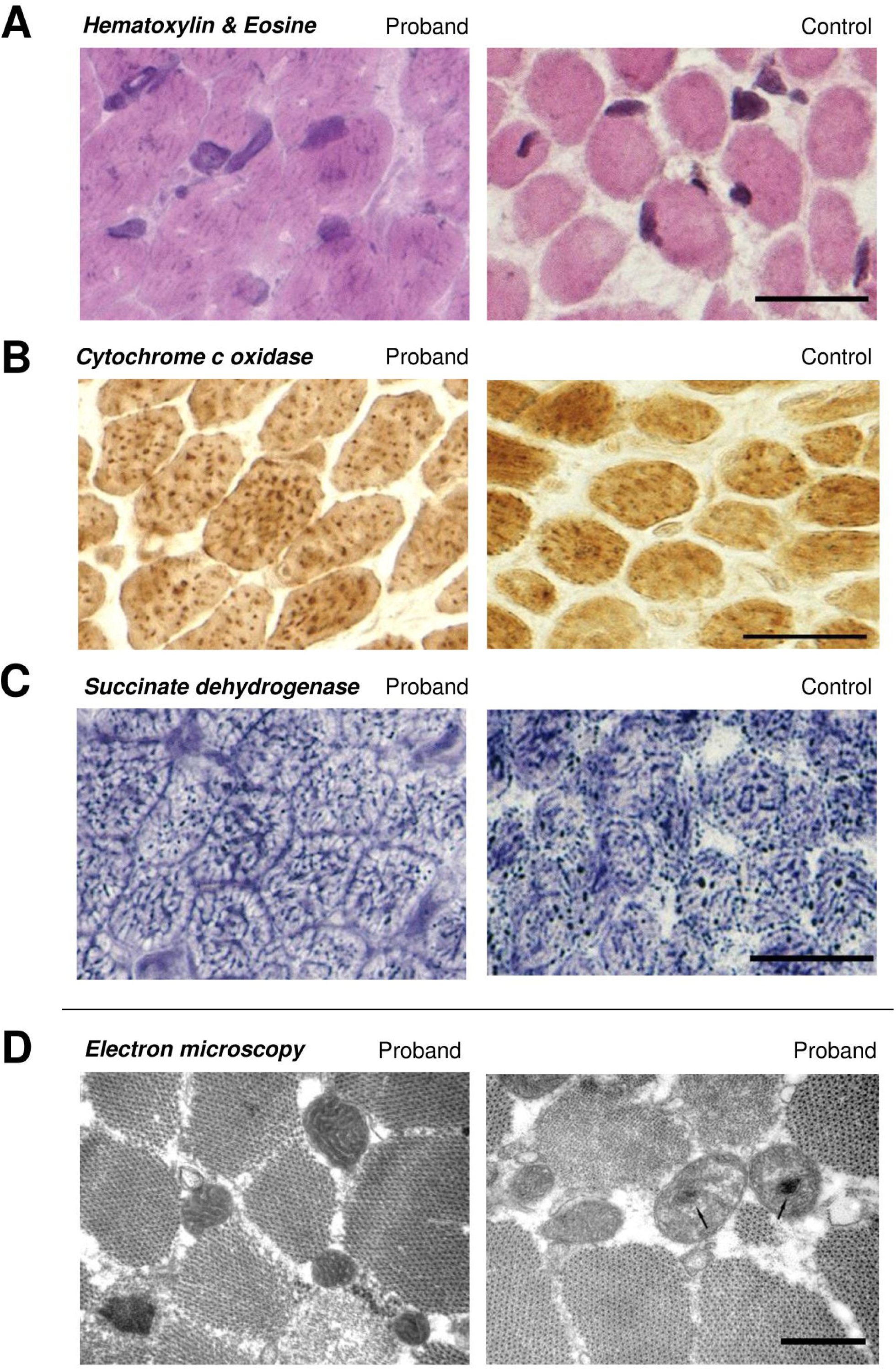
Muscle biopsy of individual 4 and an aged-matched control. The histochemical analysis revealed a prominent mitochondrial patterning. The scale bar represents 20 μm.: (A) H&E staining, in the sarcoplasm basophilic dots indicate dense and enlarged mitochondria in the patient. (B) cytochrome c oxidase staining, the mitochondria are slightly increased in size compared to the control (C) succinate dehydrogenase staining, mitochondria are evenly distributed., (D) Electron microscope (without aged-matched control), the scale bar represents 300 nm. Mitochondria show dense matrix and regular cristae. Due to the high electron density, the mitochondria appear prominent. The arrows indicate the broadened cristae by material of higher density.

In case of the variant p.(Pro194Ser) identified in individual 3, a similar destabilization is assumed. The variant is also located near the ADP binding site what may lead to steric clashes resulting from the altered sidechain geometry of Ser194 in the protein (Fig. 1B_3, 1B_4).

Individual 4 carries two compound-heterozygous variants p.(Phe341Leu) and p.(Cys6Leufs*19). The variant p.(Cys6Leufs*19) likely leads to a complete loss of the allele, likely through NMD^17^. The residue Phe341 affected by the missense variant p.(Phe341Leu) is part of a hydrophobic cluster that forms the choline binding site (Fig 1B_5). A change to Leu results in a loss of hydrophobic interactions (Fig. 1B_6), which are expected to destabilize the structure and the interaction with choline.

Individual 5 carries the homozygous start-loss variant p.(Met1?). *CHKA* has no known alternative start codons in other transcripts. The second next possible start codon occurs at amino acid position 123, potentially removing around 26% percent of the protein, and may therefore significantly impair gene expression and protein function.^18^

The variants p.(Arg141Trp) and p.(Pro194Ser) have each been observed once in a heterozygous state in the gnomAD database.^19^ For individual 4, both variants p.(Phe341Leu) and p.(Cys6Leufs*19) are absent in the gnomAD database and also no variants affecting the initiation codon have been observed (last accessed September 2021). All missense variants affect highly conserved amino acid residues and multiple *in silico* tools predict a pathogenic effect (Supplemental Table S2 and Figure S1).

### Expression of human *CHKA* in yeast

To investigate the functional significance of these variants, we assessed the effect of the individual-derived variants on *CHKA* catalytic activity. To do so we expressed the *CHKA* ORF, and the patient-derived alleles encoding these variants, from a constitutive promoter in a *S. cerevisiae* strain devoid of endogenous choline kinase activity.

Western blots showed that *CHKA* and each patient-derived variant was expressed in yeast cells at their projected molecular weight of 46 kDa and at comparable levels (Fig. 1C). Choline kinase activity for variants p.(Arg141Trp) and p.(Pro194Ser) was 20 % and 15 %, respectively, of the activity for wild type *CHKA* (Fig. 1C). These two variants were identified in homozygosity (individuals 2-4). For variant p.(Phe341Leu) catalytic activity was reduced by half. This variant is present in compound heterozygosity with a frameshift variant in the *CHKA* gene on the alternate allele (individual 4).

## Discussion

We present six individuals with bi-allelic variants in *CHKA* and establish a novel neurodevelopmental disorder of the Kennedy pathway. All affected individuals presented with a consistent phenotype of a neurodevelopmental disorder characterized by severe DD/ID, seizures starting in the first years of life, and microcephaly.

Individuals 1.1, 1.2 and 2 carry the same homozygous missense variant p.(Arg141Trp). While the phenotype was similar in almost all aspects, such as severe ID and occurrence of epileptic spasms in early infancy, individual 1.2 achieved independent walking at age three years as the only one in the cohort. What caused this difference in developmental course in this individual is unclear.

Functional testing of the variants in an *S. cerevisiae* model revealed a marked reduction of enzymatic activity ranging between 15-20% of wild-type activity for the missense variants p.(Arg141Trp) and p.(Pro194Ser). The missense variant p.(Phe341Leu) showed a reduction by half and is in a compound-heterozygous state with the frameshift variant p.(Cys6Leufs*19) that likely leads to nonsense-mediated decay mRNA transcribed from this allele. Therefore net enzyme activity is assumed to be around 25 % in this individual, which is on a comparable level to the homozygous missense variants. The functional consequence of p.(Met1?) affecting the initiation codon of *CHKA* cannot be as readily assessed.^18^ But (1) considering the consistent phenotype of individual 5 compared to the rest of the cohort, (2) the absence of an alternative start codons in other transcripts of *CHKA* and (3) the next possible methionine start codon AUG occurring at amino acid position 123, a loss-of-function mechanism and disease causality for this variant is highly likely.

Our structural *in silico* modeling of missense variants supports the assumption of reduced enzymatic activity. The variants are located near the binding sites of ATP/ADP [p.(Arg141Trp), p.(Pro194Ser)] and choline [p.(Phe341Leu)] and are therefore suggested to impair enzymatic function through structural changes or destabilization of these regions.

Compared to the other disorders described for genes of the Kennedy pathway, DD/ID and seizures seem particularly prominent in individuals of the *CHKA* cohort. When compared with its paralog *CHKB*, affected individuals also show a neurodevelopmental disorder, but seizures have rarely been reported. Severity of DD/ID ranges between mild to severe and recently, pathogenic variants in *CHKB* have been associated with autism spectrum disorder and atypical Rett syndrome.^5,20^ The abnormalities on muscle biopsy include muscular dystrophy as well as mitochondrial enlargement and placement in the periphery of muscle fibers. The muscular biopsy of individual 4 in this study also showed mitochondrial abnormalities with dense matrix and regular cristae, but mitochondria were evenly distributed throughout the cell. Even though CHKA and CHKB share a similar molecular structure and catalyze the same reaction in PC/PE biosynthesis, the phenotypic differences might be explained by different expression patterns throughout different tissues in the body.^21^ Nevertheless, no tissue specificity could be observed concerning mRNA expression or presence of protein in the cytosol.^22^ Most pathogenic variants known for *CHKB* are truncating variants leading to a complete loss of enzymatic function. In animal models, homozygous *Chkb(-/-)* mice presented with progressive muscular weakness similarly observed for the human phenotype. In comparison, *Chka(-/-)* is embryonically lethal in mice implying complete loss of CHKA activity is not compatible with vertebrate life. *Chka(+/-)* mice showed a reduction of choline kinase activity of approximately 30% but appeared to be otherwise without phenotypic abnormalities, which reinforces the assumption of recessive inheritance.^23^

The phenotypic spectrum of the *CHKA* cohort also resembles, in part, to the other disorders of the Kennedy pathway. The overlap between the phenotypes associated with *CHKA, SELENOI*, and *PCYT2* comprises DD/ID, microcephaly, short stature, visual impairment, seizures, hyperreflexia, abnormalities of muscle tone, and movement disorder (Supplemental Table 3).^8,9,24–26^ Particularly noteworthy are the eye abnormalities observed in individual 3: nystagmus, diffuse retinal pigmentary epithelium, severe conduction dysfunction and moderate retinal dysfunction in both eyes. A retinal phenotype of cone-rod dystrophy and macular pigmentary changes was described for *PCYT1A* and *SELENOI*.^7,8^

Taken together, our findings establish bi-allelic variants in *CHKA* as a novel cause of a neurodevelopmental disorder with epilepsy and microcephaly adding to the description of genetic disorders associated with the Kennedy pathway.

## Data Availability

All data produced in the present work are contained in the manuscript.

## Abbreviations

ACMG: American College of Medical Genetics
ADP: Adenosine diphosphate
cMRI: cranial MRI
COX: cytochrome c oxidase
DD/ID: developmental delay/intellectual disability
ES: exome sequencing
H&E: hematoxylin and eosin
NMD: nonsense-mediated mRNA decay
OFC: occipitofrontal circumference
ORF: open reading frame
PC: phosphatidylcholine
PE: phosphatidylethanolamine
RPE: retinal pigment epithelium
SDH: succinate dehydrogenase

## Acknowledgements

We thank all families described in this study for participating.

## Supplementary material

Supplemental data in a separate file.

