## Supplemental Data for "Bi-allelic variants in *CHKA* cause a neurodevelopmental disorder with epilepsy and microcephaly"

Chiara Klöckner, J. Pedro Fernandez Murray, Mahtab Tavasoli, Heinrich Sticht, Gisela Stoltenburg-Didinger, Leila Motlagh Scholle, Somayeh Bakhtiari, Michael C. Kruer, Hossein Darvish, Saghar Ghasemi Firouzabadi, Alex Pagnozzi, Anju Shukla, Katta Mohan Girisha, Dhanya Lakshmi Narayanan, Parneet Kaur, Reza Maroofian, Maha S. Zaki, Mahmoud M. Noureldeen, Andreas Merkenschlager, Janina Gburek-Augustat, Elisa Cali, Selina H. Banu, Kamrun Nahar, Stephanie Efthymiou, Henry Houlden, Rami Abou Jamra, Jason Williams, Christopher R. McMaster, Konrad Platzer

Content

### Supplemental Methods 1

*Individuals 1.1 and 1.2*

Whole exome sequencing was performed for the parents and the two affected children at the Yale Center for Genome Analysis (YCGA). Genomic DNA was extracted from blood and captured using IDT xGen exome kit which underwent illumina sequencing.

Sequenced reads were mapped to the reference genome h19 using BWA-MEM and processed by two GATK based pipelines at Phoenix Children’s Hospital and Yale School of Medicine. Rare single nucleotide variants (SNVs) and Indels were selected in de novo, recessive, and X-linked modes of inheritance, based on Exome Aggregation Consortium v3 (ExAC), Exome Variant Server (EVS) and 1000 Genomes databases. The deleterious variants were selected based on the “CADD score >=20 or MetaSVM=D” criteria. Confirmed variants by WES were validated using sanger sequencing.

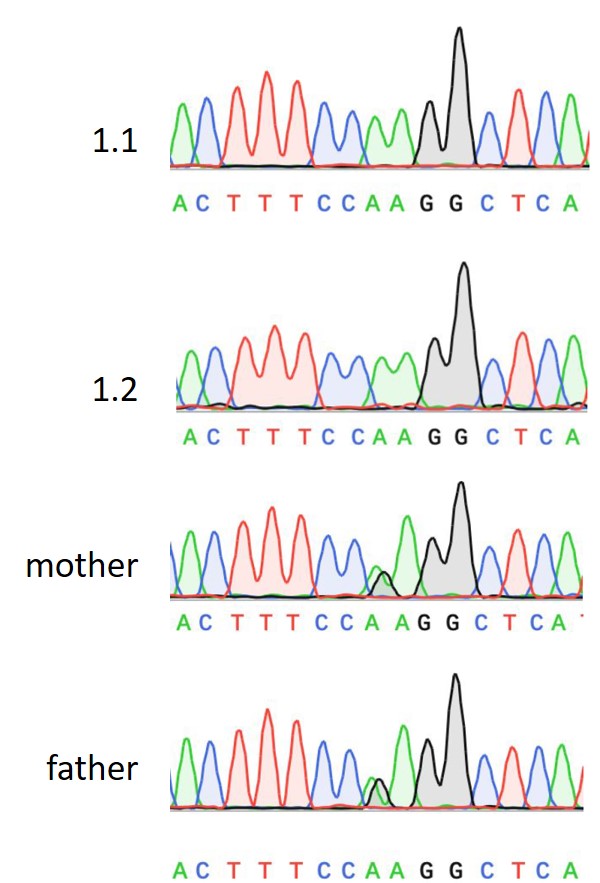

Sanger sequencing of variant in CHKA c.421C>T, p.(Arg141Trp) with homozygous state in individuals 1.1. and 1.2 und in heterozygous state in the parents.

*Individual 2*

Singleton exome sequencing (ES) was performed for the index patient. Nextera Rapid Capture Exome Kit for DNA capture (Illumina, Inc. San Diego, California, USA) was used for genomic capture. NextSeq500 Sequencer and NextSeq™ 500 High Output Kit (Illumina, Inc., San Diego, CA, USA) were used for massive parallel sequencing. ES was carried out with average coverage of 100X, 95% of bases covered at a minimum of 20X with 90% sensitivity. For ES data analysis, the raw data was retrieved in FASTQ format and aligned to GRCh37 assembly using Burrows-Wheeler Aligner (v0.7.15) and our in-house pipeline based on Genome Analysis Toolkit Best Practices. This data was annotated by ANNOVAR and our in-house scripts. Validation of the variant identified by ES and bi-allelic segregation analysis were done by Sanger sequencing.

Exome sequencing analysis rendered a homozygous missense variant in c.421C>T p.(Arg141Trp) in *CHKA* (NM_001277.2). Parents were found to be carriers for this variant.

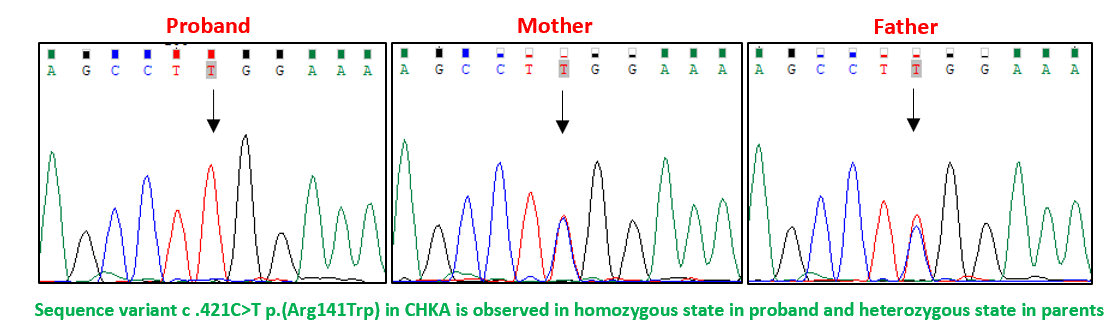

*Individuals 3 and 5*

Genomic DNA was extracted from peripheral blood samples according to standard procedures of phenol chloroform extraction. WES on each proband was performed as described elsewhere^1,2^ in Macrogen, Korea. Briefly, target enrichment was performed with 2 μg genomic DNA using the SureSelectXT Human All Exon Kit version 6 (Agilent Technologies, SantaClara, CA, USA) to generate barcoded whole-exome sequencing libraries. Libraries were sequenced on the HiSeqX platform (Illumina, San Diego, CA, USA) with 50x coverage. Quality assessment of the sequence reads was performed by generating QC statistics with FastQC (http://www.bioinformatics.bbsrc.ac.uk/projects/fastqc). Our bioinformatics filtering strategy included screening for only exonic and donor/acceptor splicing variants. In accordance with the pedigree and phenotype, priority was given to rare variants (<0.01% in public databases, including 1,000 Genomes project, NHLBI Exome Variant Server, Complete Genomics 69, and Exome Aggregation Consortium [ExAC v0.2]) that were fitting a recessive (homozygous or compound heterozygous) or a *de novo* model and/or variants in genes previously linked to developmental delay, intellectual disability, and other neurological disorders.

Individual 3:

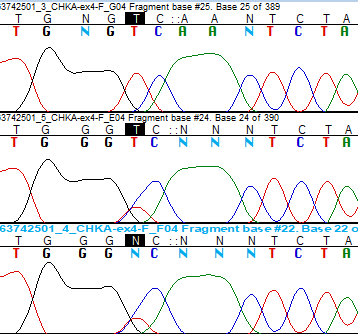

Proband

Mother

Father

Individual 5:

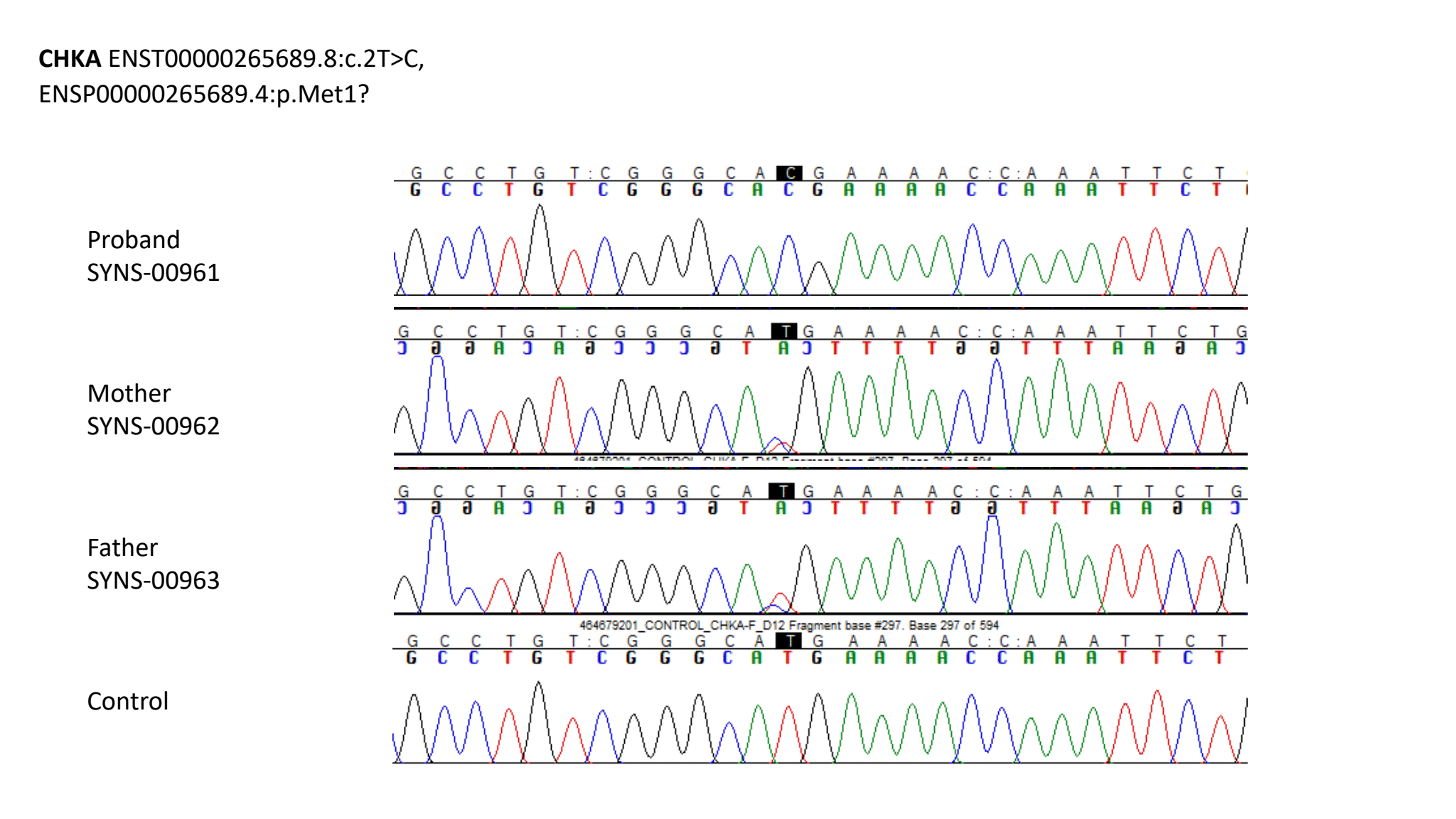

*Individual 4*

Genetic diagnostics (NGS panel diagnostics 2010, array 2010 and another NGS panel 2018 – NGS OMIM Panel) all were unremarkable) were initiated from the neuropediatric department at the age of 1 year and remained negative for clinically relevant variants. A research-based trio exome analysis was then initiated at the age of 10 years.

Acting on our instructions, the exome capture was carried out with BGI Exome kit capture (59M) and the library was then sequenced on a BGISEQ-500, paired-end 100bp, at BGI laboratory in Shenzhen, China. Analysis of the raw data was performed using the software Varfeed (Limbus, Rostock) and the variants were annotated and prioritized using the software Varvis (Limbus, Rostock). The maternally inherited missense variant NM_001277.2:c.1021T>C, p.(Phe341Leu) in exon 9 of the CHKA gene (chr11[GRCh37]:g.67833357A>G) and the paternally inherited frameshift variant NM_001277.2: c.14dup, p.(Cys6Leufs*19) in exon 1 (chr11[GRCh37]:g.67888631dup) were identified. No other candidate variants were identified to be potentially causative.

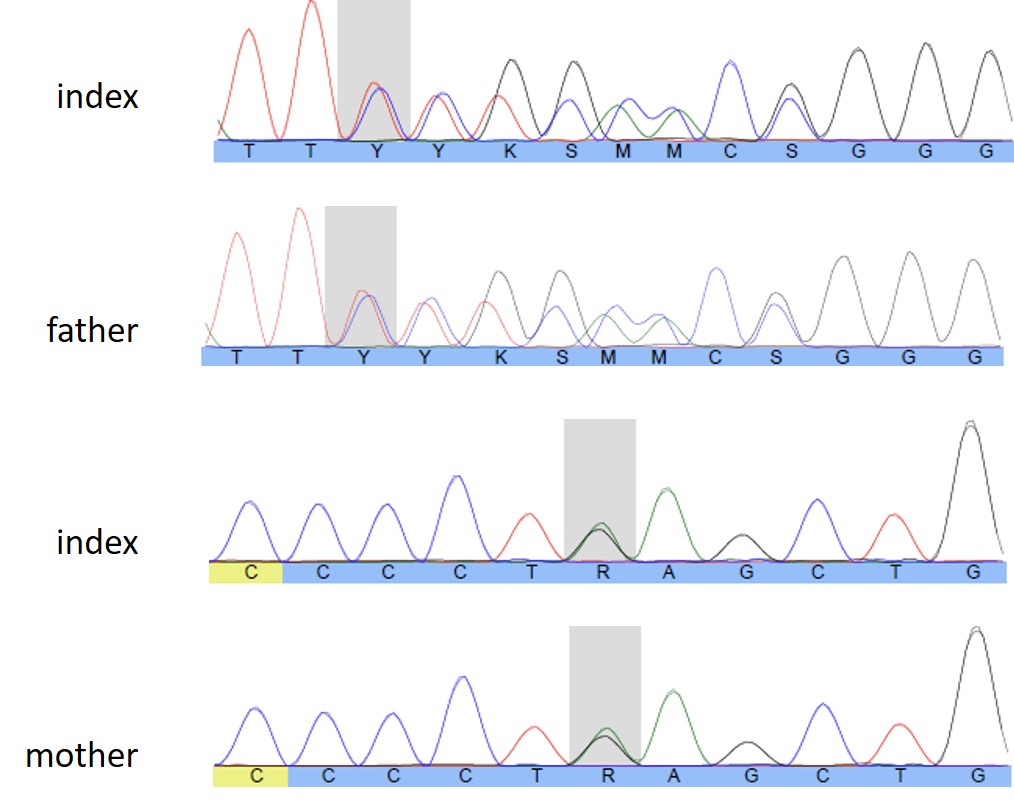

Table S1. Variant information, predicted structural effect and classification according to the ACMG criteria^3^. All variant descriptions were validated according to the HGVS nomenclature.

| **Individual** | **Chr** | **cDNA level** | **Protein level** | **Origin** | **Main structural effect(s)** | **ACMG criteria** | **Classification** |
| --- | --- | --- | --- | --- | --- | --- | --- |
| 1.1, 1.2, 2 | g.67864527G>A | c.421C>T | p.(Arg141Trp) | biparental | Impairment of ATP binding | PS3, PS4_Moderate, PM2_Supporting, PP3 | Likely pathogenic |
| 3 | g.67842234G>A | c.580C>T | p.(Pro194Ser) | biparental | Impairment of ATP binding | PS3, PM2_Supporting, PP3 | Likely pathogenic |
| 4 | g.67888631dup | c.14dup | p.(Cys6Leufs*19) | paternal | Nonsense mediated mRNA decay^4^ | PVS1, PM2_Supporting | Likely pathogenic |
| 4 | g.67833357A>G | c.1021T>C | p.(Phe341Leu) | maternal | Impairment of substrate binding | PS3, PM2_Supporting, PP3 | Likely pathogenic |
| 5 | g.67888643A>G | c.2T>C | p.(Met1?) | biparental | Unclear* | PVS1_Supporting, PM2_Supporting | Uncertain significance* |

* Although this variant is very likely disease causing, it has to be classified as of uncertain significance using the ACMG criteria. *CHKA* has no known alternative start codons in other transcripts. The second next possible start codon for methionine occurs at amino acid position 123 (removing around 26% percent of the protein) and therefore likely significantly impairs protein function.^5^ In the context of a presumed loss-of-function mechanism of pathogenic variants in *CHKA*, a disease-causing effect of variants affecting the initiation codon is likely. Furthermore, no homo- or heterozygous variants affecting the initiation codon have been observed in the gnomAD database (last accessed 09/2021).^6^

Table S2. *In silico* prediction of missense variants and conservation of affected amino acids in *CHKA*.

| **Individual** | **Chr** | **cDNA level** | **Protein level** | **CADD 1.4**^7^ | **REVEL**^8^ | **Mutation Taster2**^9^ | **M-CAP 1.3**^10^ | **Polyphen 2 v2.2.2**^11^ | **GERP++** | **Conservation** | **Allele Count in gnomAD**^6^ |
| --- | --- | --- | --- | --- | --- | --- | --- | --- | --- | --- | --- |
| 1.1, 1.2, 2 | Chr11:67864527 G>A | c.421C>T | p.(Arg141Trp) | 26.9 | 0.38 | DC | D | PrD | 5.74 | Moderate (fruitfly) | 1 |
| 3 | Chr11:67842234 G>A | c.580C>T | p.(Pro194Ser) | 28 | 0.91 | DC | D | PrD | 4.97 | High (Baker’s yeast) | 1 |
| 4 | Chr11:67833357 A>G | c.1021T>C | p.(Phe341Leu) | 29.6 | 0.76 | DC | D | PrD | 4.97 | High (Baker’s yeast) | 0 |

Conservation was evaluated considering the following species: Homo sapiens, Pan troglodytes (chimp), Rattus norvegicus (rat), Mus musculus (mouse), Canis familiaris (dog), Ornithorhynchus anatinus (platypus), Gallus gallus (chicken), Xenopus tropicalis (frog), Tetraodon nigroviridis, Danio rerio (zebrafish), Drosophila melanogaster (fruitfly), Caenorhabditis elegans (C. elegans), Saccharomyces cerevisiae (Baker’s yeast).

The color represents the probability of the variant to be damaging for each aspect in the table: red *– in silico* damaging; highly conserved amino acid; absent/low frequency in gnomAD, yellow – *in silico* mixed results; moderately conserved amino acid; low frequency in gnomAD.

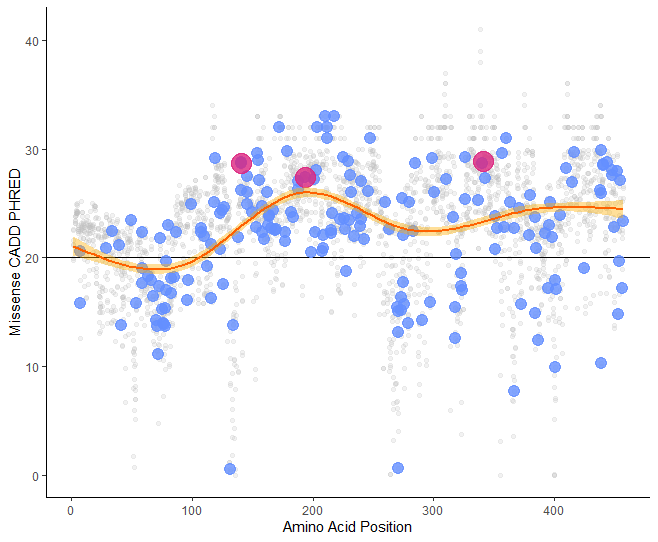

Figure S1. A generative additive model shows the values of CADD PHRED v1.4 for all possible missense variants in *CHKA* (grey points). Blue points represent variants observed in gnomAD, red points variants observed in this cohort. The black horizontal line (CADD PHRED = 20) marks the recommended cut-off for potential pathogenicity.

### Supplemental Methods 2

Muscle biopsy: 7-8-μm thin cryosections from the skeletal muscle biopsy were stained with H&E, SDH, and COX. COX-SDH double staining according to standard protocols.

Selected specimens of the muscle biopsy were fixed in 3% glutaraldehyde for subsequent ultrastructural investigation, postfixed with OsO4 and embedded in Araldite epoxy resins that polymerized for 16h at 60°C. Ultrathin sections were cut at 50-70 nm, mounted on copper nets, double-contrasted with uranyl acetate and lead citrate and analyzed with a "Zeiss EM 906" at up to 40.000fold magnification at on operating voltage of 80 kV.

### Case reports

*Individuals 1.1 and 1.2*

This case report has been removed from the preprint version to comply with medrxiv policy. Please see the published version or contact the authors if you are interested in this information.

*Individual 2*

This case report has been removed from the preprint version to comply with medrxiv policy. Please see the published version or contact the authors if you are interested in this information.

*Individual 3*

This case report has been removed from the preprint version to comply with medrxiv policy. Please see the published version or contact the authors if you are interested in this information.

*Individual 4*

This case report has been removed from the preprint version to comply with medrxiv policy. Please see the published version or contact the authors if you are interested in this information.

*Individual 5*

This case report has been removed from the preprint version to comply with medrxiv policy. Please see the published version or contact the authors if you are interested in this information.

**MRI Images**

*Individual 1.1*

These images have been removed from the preprint version to comply with medrxiv policy. Please see the published version or contact the authors if you are interested in this information.

*Individual 2*

These images have been removed from the preprint version to comply with medrxiv policy. Please see the published version or contact the authors if you are interested in this information.

*Individual 4*

These images have been removed from the preprint version to comply with medrxiv policy. Please see the published version or contact the authors if you are interested in this information.

Table S3. Overlap of phenotypes in the Kennedy pathway^12–19^

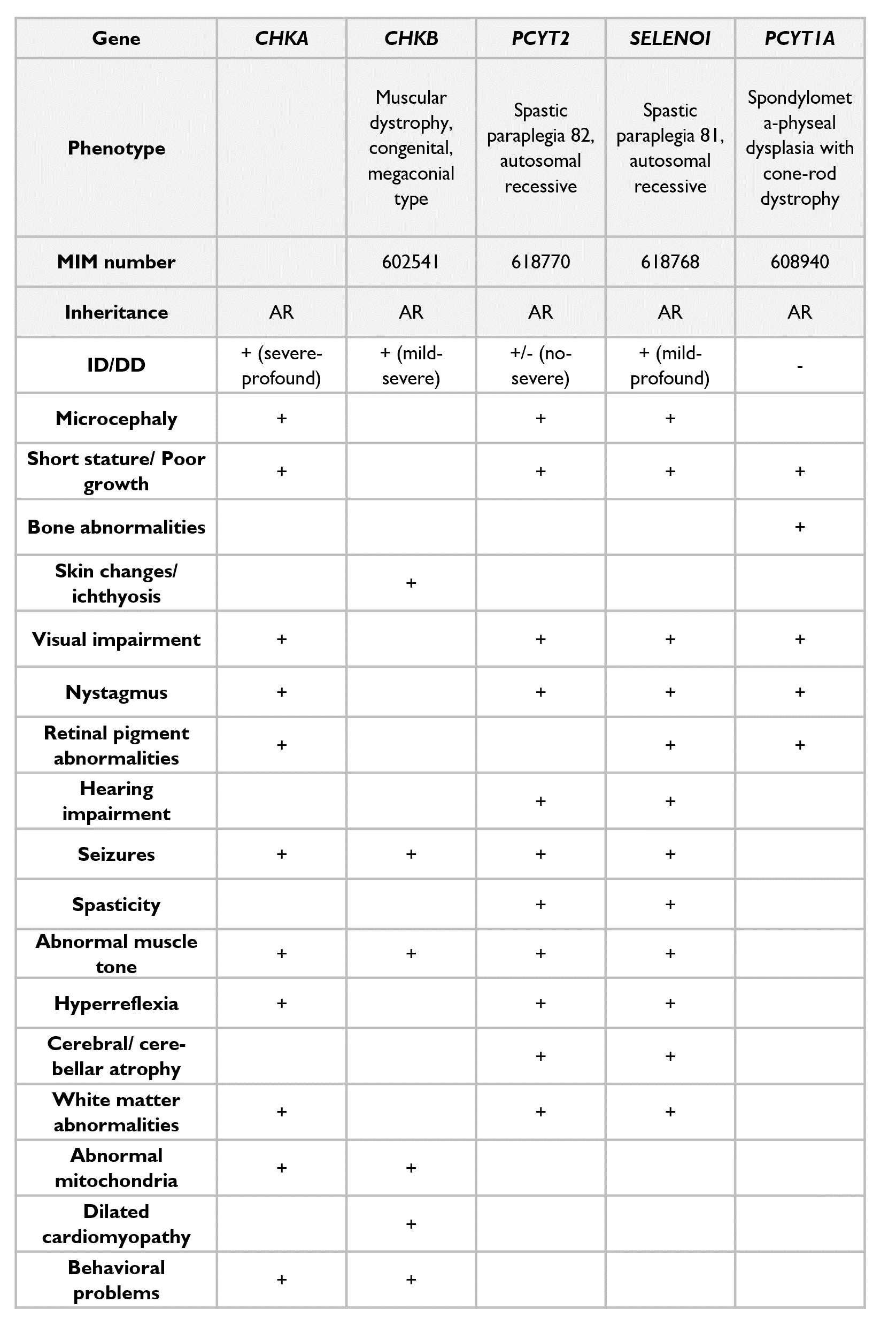
